# Transmission Clusters of Hepatitis C Virus among Public Health and Correctional Settings in Wisconsin, 2016-2017

**DOI:** 10.1101/2020.06.19.20134148

**Authors:** Karli R. Hochstatter, Damien C. Tully, Karen A. Power, Ruth Koepke, Wajiha Z. Akhtar, Audrey F. Prieve, Thomas Whyte, David J. Bean, David W. Seal, Todd M. Allen, Ryan P. Westergaard

## Abstract

Ending the hepatitis C virus (HCV) epidemic requires stopping transmission among networks of people who inject drugs (PWID). Identification of transmission networks through the application of genomic epidemiology may inform community response models that can quickly interrupt transmission. We retrospectively identified HCV RNA-positive specimens corresponding to 459 individuals tested in public health settings, including correctional facilities and syringe service programs, in Wisconsin from 2016-2017. Next-generation sequencing of HCV was conducted and analyzed for phylogenetic linkage using the CDC’s Global Hepatitis Outbreak Surveillance Technology platform. Transmission network analysis showed that 126 individuals were linked across 42 clusters (range: 2-11 individuals per cluster). Phylogenetic clustering was higher in rural communities and associated with female gender and younger age among rural residents. These data highlight that the increasing rurality of opioid injection use and HCV transmission among young PWID could be better supported by the expansion of molecular-based surveillance strategies to reduce transmission.

**Article Summary Line:** Integrating existing public health surveillance and molecular analyses with Global Hepatitis Outbreak Surveillance Technology allows for the identification and characterization of growing HCV transmission clusters among key populations in Wisconsin.

## INTRODUCTION

Sharp increases in hepatitis C virus (HCV) infection have been observed in the United States, where an estimated 2.4 million persons are living with chronic infection *(1)*. In 2013, approximately 19,368 persons died from HCV-related complications, exceeding the number of deaths from all other nationally notifiable infectious diseases combined *(2)*. A 2-fold increase in the prevalence of HCV has occurred between 2004 and 2014, a direct result of the opioid epidemic and associated increases in the sharing of contaminated injection drug use equipment *(3)*. As the intersecting epidemics of opioid injection and infectious diseases are complex and dynamic, implementing community-specific comprehensive prevention services has remained challenging. Reliable strategies to identify individuals and communities most vulnerable are essential for the development of community response models able to deliver evidence-based prevention services in time to prevent worsening of these synergizing epidemics.

Epidemiologists and public health experts are increasingly utilizing molecular-based surveillance techniques to identify and control emerging outbreaks *(4–7)*. For example, the US Centers for Disease Control and Prevention (CDC) has recently scaled up the use of molecular HIV surveillance with pilot projects in 27 states towards an “Ending the HIV epidemic” initiative *(8)*. Expanded use of molecular surveillance has the potential to significantly inform HCV treatment and prevention efforts. However, application of such programs for HCV surveillance has lagged despite the potential importance of such approaches in disrupting HCV transmission. Global Hepatitis Outbreak and Surveillance Technology (GHOST) is a public health tool developed by CDC that uses next generation sequencing methods for molecular HCV surveillance and outbreak investigation *(9)*. Analysis of next-generation sequencing data for viruses is complex and particularly challenging and requires significant expertise in bioinformatics and phylogenetics. GHOST integrates a suite of computational tools to accurately detect possible HCV transmission clusters from next generation sequencing data in a simple fashion regardless of the user’s level of expertise. The cloud-based system provides the user with detailed reports on the likelihood of linked transmission events within a cohort of HCV-infected individuals, thus providing an opportunity for the rapid identification of existing or growing HCV transmission clusters and an improved response by state and local public health departments.

Wisconsin is a state in the U.S. with a population of 5.8 million residents. Between 2016 and 2017, the rate of opioid overdose in Wisconsin increased 109%, representing the steepest increase observed in any U.S. state, and nearly 3 times the average national increase over that time period *(10)*. Similar to what has been observed nationally, this sharp increase in opioid use is accompanied by substantial increases in the incidence of HCV. Between 2011 and 2015, approximately 3,000 new HCV diagnoses were reported annually. The rate of new PCR-confirmed HCV diagnoses among people aged 15-29 more than doubled during that time period, from 40 to 87 cases per 100,000 population, as a result of recent injection drug use *(11)*.

In this study, we integrate existing public heath surveillance and molecular analyses with GHOST to identify putative HCV transmission clusters among persons likely infected through injection drug use during a period of expanded HCV transmission *(12)* and investigate the network characteristics among this high-risk group.

## MATERIALS AND METHODS

### Study Setting

All HCV-positive reports in Wisconsin are routinely reported to the Wisconsin Electronic Disease Surveillance System (WEDSS), a secure, web-based health information system used for the reporting, investigation, and surveillance of communicable diseases in Wisconsin. Blood samples collected for HCV RNA confirmatory testing at sites supported by the Wisconsin Division of Public Health (DPH) are processed at the Wisconsin State Laboratory of Hygiene and stored for up to 5 years. Approximately 15% of all HCV cases reported to WEDSS represent individuals who underwent fee-exempt HCV RNA confirmatory testing through the State Laboratory; these instances typically reflect testing done in public health settings such as syringe service programs and correctional facilities, and not traditional health care settings. This results in a cohort that is enriched for younger people with a history of injection drug use.

### Study Population

Individuals confirmed to have an HCV RNA-positive sample analyzed at the State Laboratory and reported to WEDSS for the first time during 2016-17 were identified using two methods. First, we identified 241 individuals residing in rural catchment areas. Counties included in the rural catchment area were selected based on participation in an ongoing federally funded research program, and were classified as rural because they were served by one of the statewide syringe service program’s six rural offices. Secondly, we identified an additional cohort of 218 individuals outside of the targeted rural area who were considered likely to represent recent or acute infections because they had either (1) presented as acute HCV when reported to WEDSS or (2) were age 15-39 at diagnosis and had a HCV viral load >1,000,000 IU/L. This second cohort, which included counties served by one of the syringe service program’s four urban offices as well as correctional populations, was included to improve network completeness and compare the extent of clustering between rural and non-rural populations.

### Specimen Processing

Per standard protocol, serum remaining after HCV antibody and RNA PCR testing is completed is stored at the State Laboratory at −80°C. Specimens corresponding to the HCV RNA-positive individuals identified in WEDSS were retrieved from the freezer and overnight shipped on dry ice to the Ragon Institute of MGH, MIT and Harvard for viral sequencing.

### Nucleic acid extraction and PCR amplification

RNA was isolated from 140 μl of plasma using the QIAamp Viral RNA Mini Kit (Qiagen, Hilden, Germany). A one step RT-PCR reaction was performed on all samples to amplify a segment at the E1/E2 junction of the HCV genome which contains the hypervariable region 1 (HVR1) *(13)*. This region was chosen for its high variability and its ability to reliably detect transmission events in outbreak settings *(14)*. The first round RT-PCR consisted of an Illumina adapter specific portion, a sample specific barcode segment, and an HCV HVR specific primer segment, F1-GTGACTGGAGTTCAGACGTGTGCTCTTCCGATCT-NNNNNNNNNN-GGA-TAT-GAT-GAT-GAA-CTG-GT and R1-ACA-CTC-TTT-CCC-TAC-ACG-ACG-CTC-TTC-CGA-TCT-NNNNNNNNNN-ATG-TGC-CAG-CTG-CCG-TTG-GTG-T at a final concentration of 4 pM amplified using Superscript III RT/Platinum Taq DNA Polymerase High Fidelity with the following conditions: cDNA synthesis for 30 minutes at 55°C, followed by heat denaturation at 95°C for 2 minutes, the PCR amplification conditions were 40 cycles of denaturation (94°C for 10 seconds), annealing (55°C for 10 seconds) and extension (68°C for 10 seconds) with a final extension at 68°C for 5 minutes. Amplified products were run on a 1% agarose gel and either PCR purified with the Qiaquck PCR purification kit (Qiagen) or gel extracted and purified using the PureLink quick gel extraction kit (Invitrogen). A second round limited cycle PCR (94°C for 2 minutes, (94°C for 15 sec; 55°C for 30 sec; 68°C for 30 sec) x 8 cycles, 68°C for 5 minutes) is performed to add barcode specific indexes and sequencing specific adapters and primers to each sample to allow for multiplexing as well as internal controls for cross-contamination. Negative controls were introduced at each stage of the procedure and all PCR procedures were performed under PCR clean room conditions using established protocols. Indexed samples are 0.7X SPRI purified two times to remove excess primer dimer and short fragments that can interfere with the sequencing process.

### Illumina deep sequencing and analysis

Resulting PCR amplicons were quantified using the Picogreen kit (Invitrogen, Carlsbad, CA) on a Fluorometer ST (Promega, Madison, WI) with the integrity of the fragment evaluated using a Bioanalyzer 2100 (Agilent, Santa Clara, CA). Samples were pooled and sequenced on an Illumina MiSeq platform using a 2 × 250 bp V2 Nano reagent kit. In general, a sequence library consisted of between 8-16 specimens including one negative control for every 7 serum specimens. Paired end reads were subject to stringent cleaning and quality control criteria as outlined previously *(15–17)*. Duplicate reads were removed using fastuniq v1.1 *(18)* and quality trimmed using trimmomatic v0.36 *(19)*. Viral contigs were generated using Vicuna v1.1 *(20)* and a *de novo* consensus assembly generated using V-FAT v1.1.

### Phylogenetic reconstruction

Consensus sequences were aligned using MEGA v6.0 *(21)* and IQ-TREE v1.6 *(22)* was used to construct a maximum likelihood phylogenetic tree with 1,000 ultrafast bootstrap replicates *(23)*.

### HCV transmission network analysis

Illumina paired end reads were uploaded to GHOST where they were subjected to automatic quality control that rejects data depending on the magnitude of the quality problem and warns and guides users in resolving the issue. Further methodological details on the quality filtering can be found elsewhere *(9,24)*. Transmission links that represent the genetic similarity among viral populations from infected individuals were examined by the transmission analysis module of GHOST. In each case the intra-host populations were compared between infected individuals and the genetic distance, defined as the Hamming distance, between their closest haplotypes was calculated. If the genetic distance was smaller than an empirically defined genetic distance threshold of 3.77% then individuals are considered to be genetically related and form a transmission cluster *(14)*. To further analyze the genetic relationships in each cluster and to visualize the nature of intra-host variation, k-step networks of intra-host HCV HVR1 variants were built as previously described *(9)*.

### Data Collection

Variables routinely reported to WEDSS for individuals who test HCV-positive include age, gender, race/ethnicity, HCV-positive antibody and RNA test date(s), and testing site(s). Additionally, individuals tested by a community-based multisite syringe service program, operated by 10 offices across the state and a mobile van, provide risk information through the syringe service program’s standard HCV testing procedures. Reporting of risk information is not mandatory. When possible, local health department staff gather risk information from the health care provider or directly from the patient and enter the risk information into WEDSS. HCV cases originally reported from state correctional facilities are not interviewed by local public health and typically do not have risk information available. For individuals with risk information available, we assessed whether they ever engaged in injection drug use, shared drug injection equipment, are a male who has sex with other males (MSM) or were ever incarcerated. Persons were considered to be incarcerated if they had any HCV test result reported to WEDSS that was conducted at a state correctional facility or if the person reported ever being incarcerated on risk information forms. Because availability of risk information depends on the type of facility where the individual was tested, we present demographic characteristics and risk behaviors by type of testing facility: community-based organization, corrections, local health department, or other public venues.

Data were collected under a waiver of informed consent. This study was approved by the University of Wisconsin’s Health Sciences Institutional Review Board and the Massachusetts General Hospital Institutional Review Board. Data Use Agreements and a Materials Transfer Agreement were established between the University of Wisconsin, DPH, the State Laboratory, and the Ragon Institute of MGH, MIT and Harvard.

### Statistical Analyses

Chi-square, Fisher’s exact, t-test, and analysis of variance (ANOVA) were conducted using StataSE version 16 (Statacorp, College Station, TX) to compare clustering by demographics and risk behaviors. Because sampling techniques differed in rural and non-rural catchment areas and the characteristics assessed were strongly determined by which catchment area they were in, we compared individuals who clustered to those who did not cluster stratified by catchment area. We also compared characteristics between rural catchment area-only clusters, non-rural catchment area-only clusters, and clusters that contained individuals from both groups. Statistical significance was determined using α=.05.

## RESULTS

### Study Sample

Between 2016 and 2017, 459 individuals tested HCV RNA-positive in public health settings in Wisconsin for the first time. Of these, 424 (92.4%) had sufficient (approximately 200ul or more) residual serum specimens stored in the State Laboratory freezer. All 424 specimens were shipped for viral sequencing. Of these, 379 of 424 specimens (89.4%) were successfully amplified, sequenced, and passed GHOST quality control metrics. Among the specimens that failed, 23 (5.4%) failed PCR and 22 (5.2%) failed GHOST quality control metrics. After quality control the median number of error-corrected reads per individual was 16,740 (IQR: 13,302 – 18,262) while the median number of haplotypes was 3,322 (IQR: 2,479 – 4,345).

### Patient Demographic Characteristics and Risk Behaviors

Among the 379 individuals whose specimens were successfully analyzed by GHOST, 125 (33.0%) first tested positive at a community-based organization, 154 (40.6%) in a correctional facility, 40 (10.6%) at a local health department, and 60 (15.8%) in a different public health setting (**Table 1**). The study population was primarily non-Hispanic white (85.0%), aged 18-39 years (90.8%), and male (75.5%). Self-report of injection drug use was documented among 177 (46.7%) individuals, and of these, 145 (81.9%) self-reported ever sharing injection equipment. MSM was reported for 8 (2.1%) individuals. The majority of the study population has experienced incarceration (88.1%).

**Table 1:**
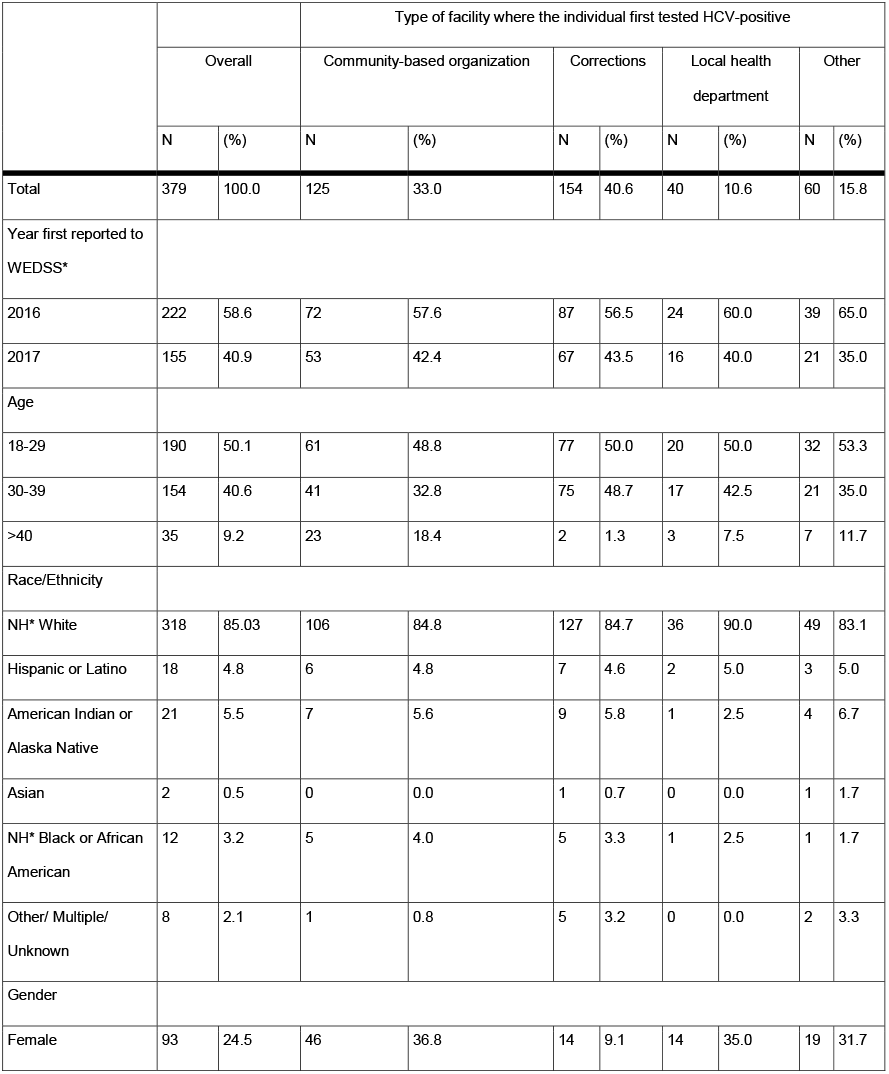
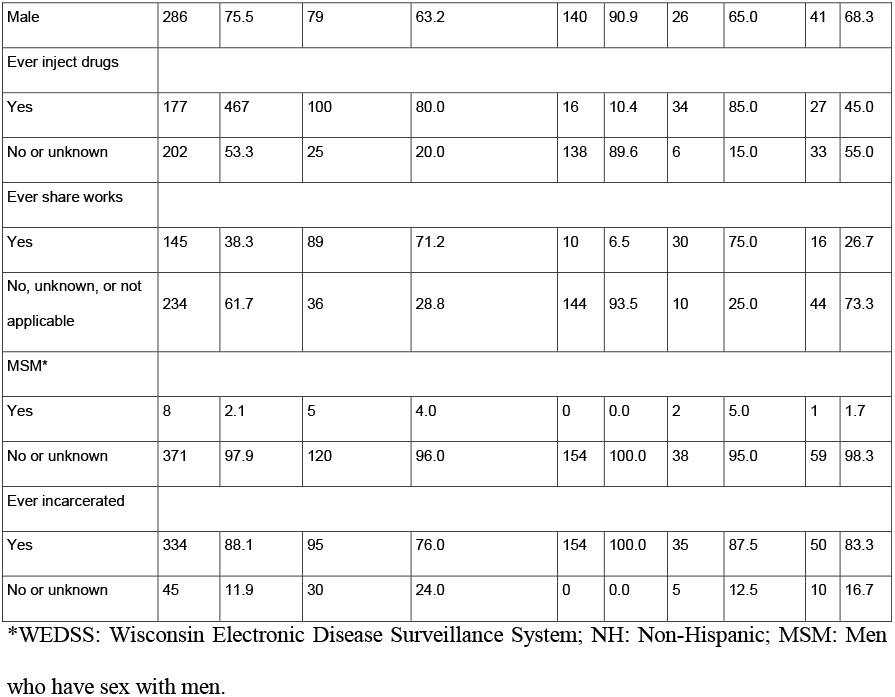
Demographic and risk factor information, by type of testing facility

Among the 379 individuals, 171 (45.1%) resided in the rural catchment area, of which 67 (39.2%) clustered, and 208 (54.9%) resided outside of the rural catchmant area or in correctional facilities, of which 59 (28.4%) clustered. Among the 171 individuals in the rural catchment area, females were significantly more likely to cluster than males (49% vs 33%) *(P*=0.04), and individuals who clustered were significantly younger (mean age: 28.7) than individuals who did not cluster (mean age: 34.1) *(P*=0.0001). There were no statisically significant differences between those who clustered and those who did not cluster in non-rural catchment areas.

### Phylogenetic analysis

HCV strains were genetically characterized using HVR1 consensus sequences derived from all 379 individuals. Phylogenetic analysis demonstrated a predominance of genotype 1a (n=255, 67.3%) and 3a (n=88, 23.2%) infections, followed by 2b (n=22, 5.8%), 1b (n=9, 2.4%), 2a (n=4, 1.1%) and 4a (n=1, 0.3%) (**Figure 1**). Within each of the major subtypes, sequences were composed of multiple lineages with the largest HVR1 cluster involving over half (54%) of all subtype 1a sequences generated. These observations suggest multiple independent HCV introductions into this population in the past.

**Figure 1.**
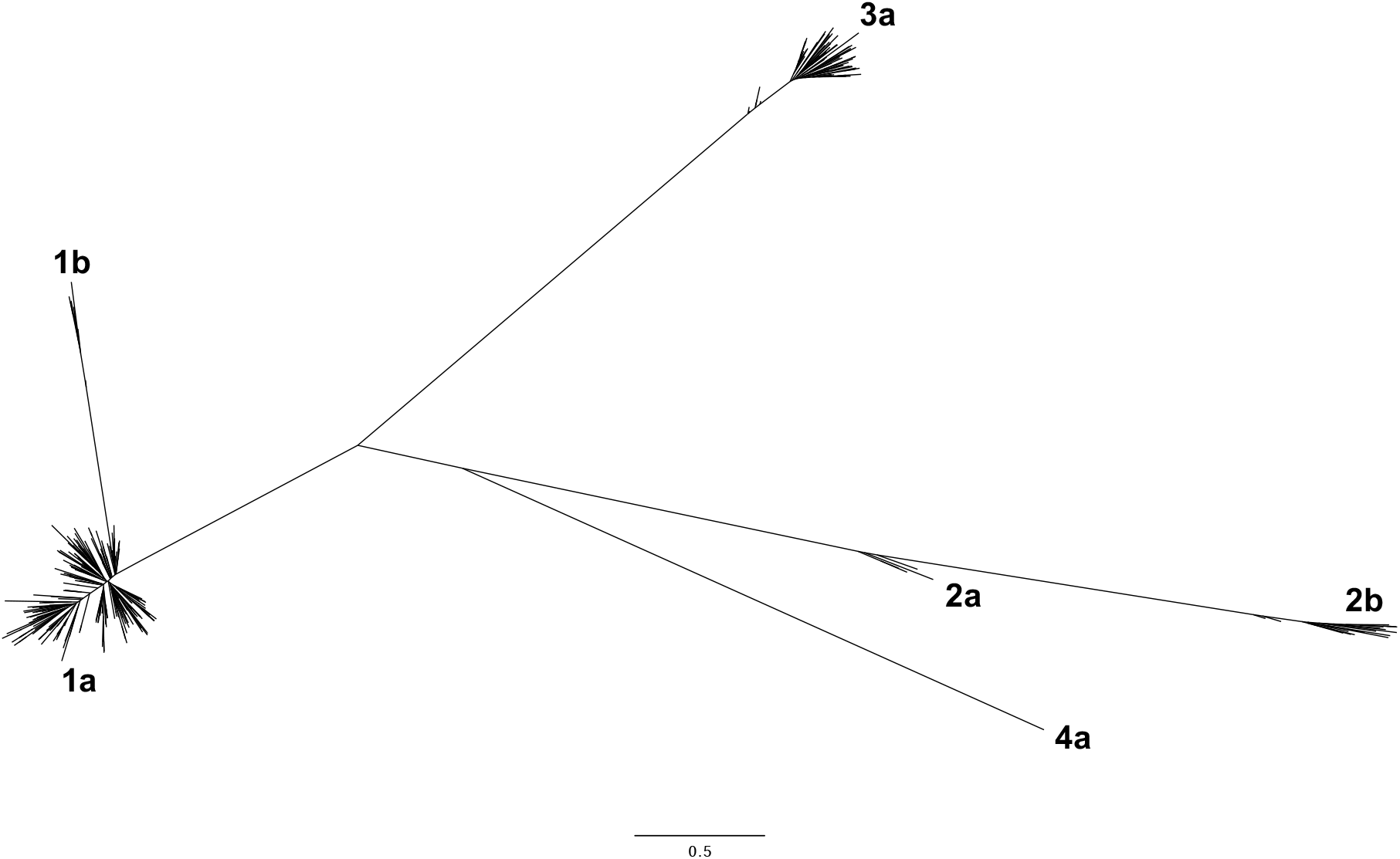
Maximum likelihood phylogenetic tree of HCV HVR1 consensus sequences. 379 consensus sequences from Wisconsin are depicted with genotypes labelled.

### HCV Transmission Linkages

GHOST detected 42 clusters comprising 126 individuals for an overall clustering rate of 33% (**Figure 2**). The size of clusters ranged from 2 to 11 individuals. The transmission networks were composed of mostly dyads (n=23, 54.8%), followed by groups of 3 (n=9, 21.4%), 4 (n=3, 9%) and 5 (n=6, 14.3%). The largest cluster involved 11 individuals, all infected with genotype 3a, with two individuals first reported to WEDSS in 2016 and nine from 2017. Five of the 11 first tested HCV-positive by the same local health department, and three first tested HCV-positive at the same syringe service clinic. Evidence of past injection drug use was available for 7 individuals, 8 were male, and all 11 were non-Hispanic white with a history of incarceration. Among the 42 clusters identified, 12 were comprised only of individuals not residing in the rural catchment area (n=27), 12 were comprised only of individuals residing in the rural catchment area (n=34), and 18 contained individuals from both groups (n=65). Rural catchment area-only clusters were more likely to have a higher percentage of females. No other significant differences between rural, non-rural, and mixed clusters were found.

**Figure 2.**
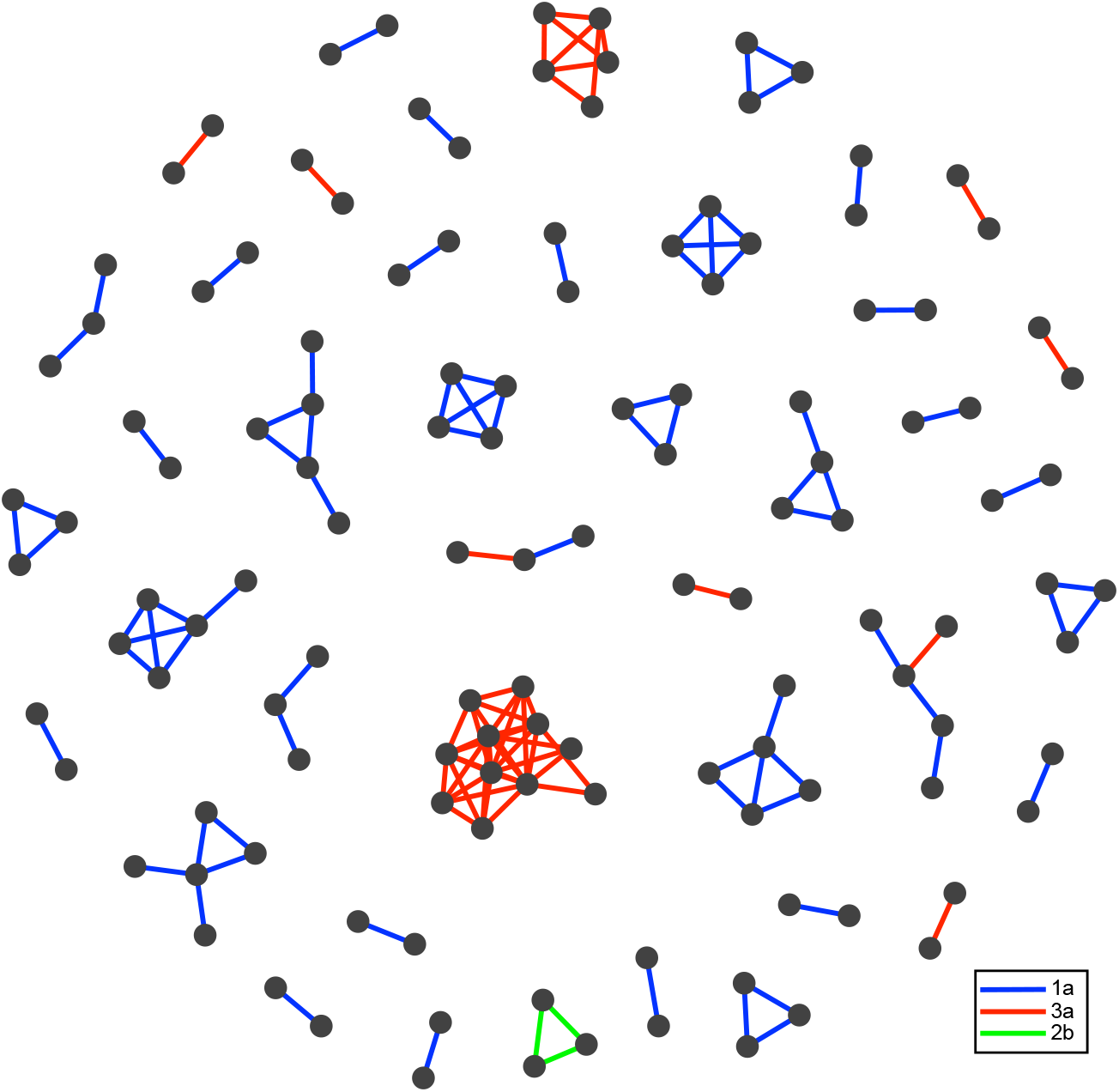
HCV transmission network of clusters identified by GHOST. Each node represents a HCV-infected individual for which HCV sequence data was generated. A transmission link is denoted as a line connecting individuals where the minimal hamming distance between sequences is smaller than the previously validated genetic threshold of 3.77%. Lines connecting clusters are colored according to genotype with a blue, red and green line corresponding to genotypes 1a, 3a and 2b cluster respectively.

### Intra-host genetic variation within transmission clusters

GHOST analysis of the intra-host HVR1 variants in 379 individuals revealed five individuals (1.3%) were infected with multiple HCV strains (**Table 2**). In all cases, genotype 1a was the major strain, with two individuals harboring minor variants of genotype 3a and genotypes 2a, 2b and 6f also found. To further understand the nature of HCV transmission across clusters, we examined the population structure of HVR1 variants to address whether the same viral variant was shared among HCV infected individuals as previously described (14). While it is not possible to illustrate the k-step networks for each sample, we highlight here representative examples of clusters. One cluster comprises three individuals (subjects 372, 338 and 362) with both subjects 338 and 362 harboring little intra-host genetic variation and sharing 19 viral variants (modified Hamming distance = 0) (**Figure 3a**). The third individual, subject 372, is infected with many diverse variants with a single subpopulation that is genetically similar to those variants found in subjects 338 (**Figure 3b**) and 362 (**Figure 3c**). Another representative cluster comprises a simple dyad of subjects 84 and 86 which share a variant (modified Hamming distance = 0.37) that only has a minor difference between each other (**Figure 4a**). In contrast, the cluster of subject 281 and 367 shares a more distantly related variant (modified Hamming distance = 3.18) (**Figure 4b**). Taken together, this reveals a complex picture of variable and extensive interconnectedness between variants with only a minority of individuals (n=14) sharing the same viral variant within a transmission cluster.

**Table 2.**
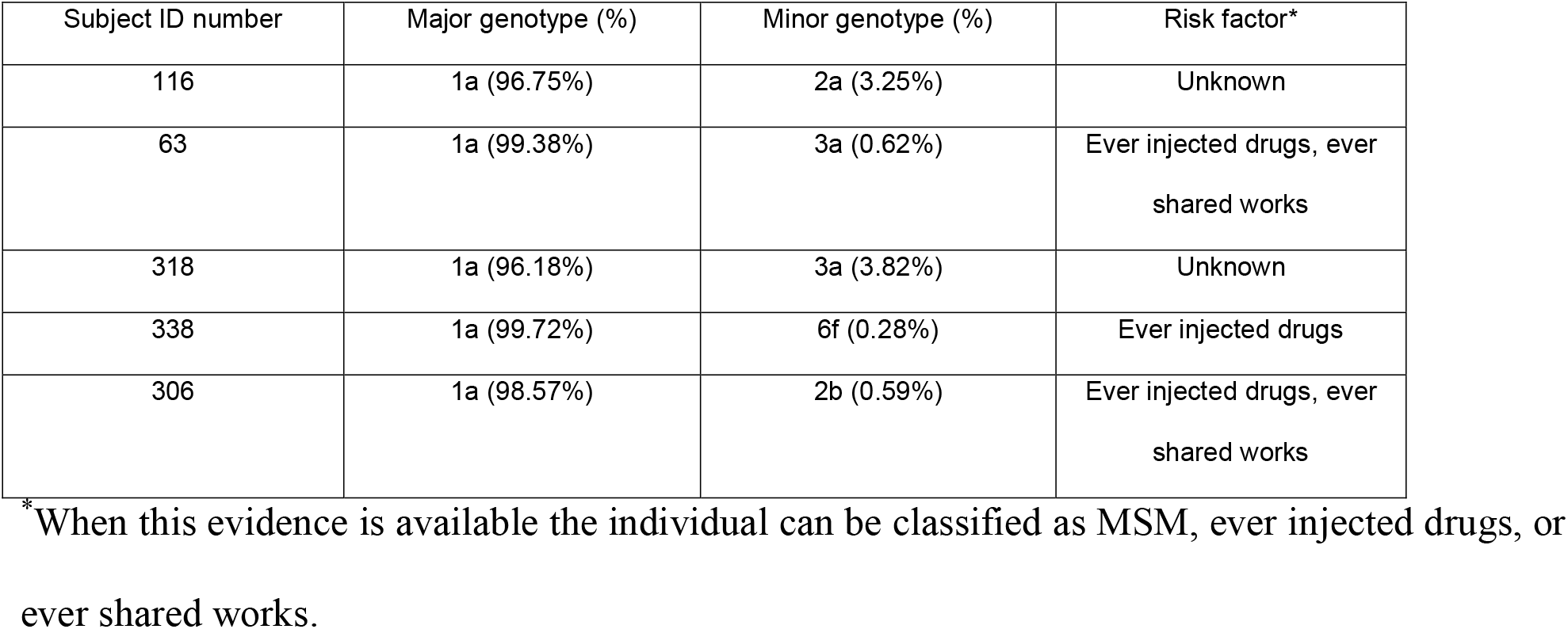
Characteristics of samples infected with mixed HCV genotypes

**Figure 3.**
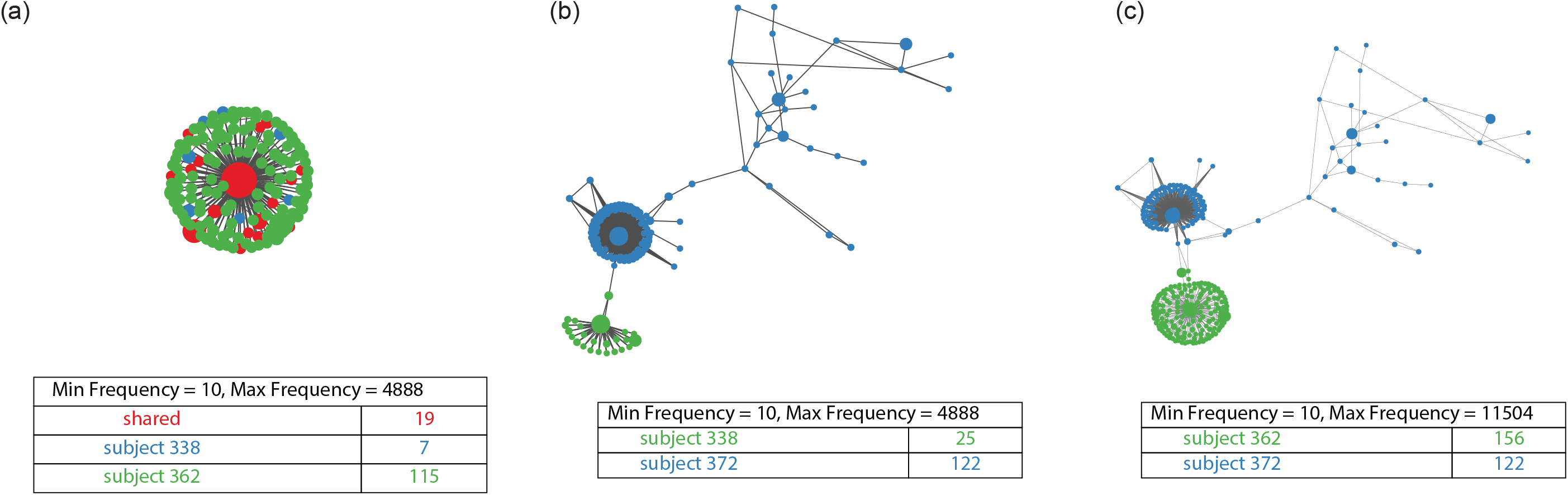
Intra-host genetic heterogeneity within transmission clusters. K-step network contains all possible minimum spanning trees and allows the efficient visualization of genetic relatedness among all intra-host HVR1 variants for **(a)** Subjects 338 and 362 **(b)** Subjects 338 and 372 and **(c)** Subjects 362 and 372. Each node represents an HCV sequence and the color of the node corresponds to the sample of origin: red if the variant was found in both samples, green if it was only found in the first sample and blue if it was only found in the second sample. The node size is based on frequency of the HVR1 variant and edge length is proportional to the modified Hamming distance (does not count positions with insertions or deletions as differences).

**Figure 4.**
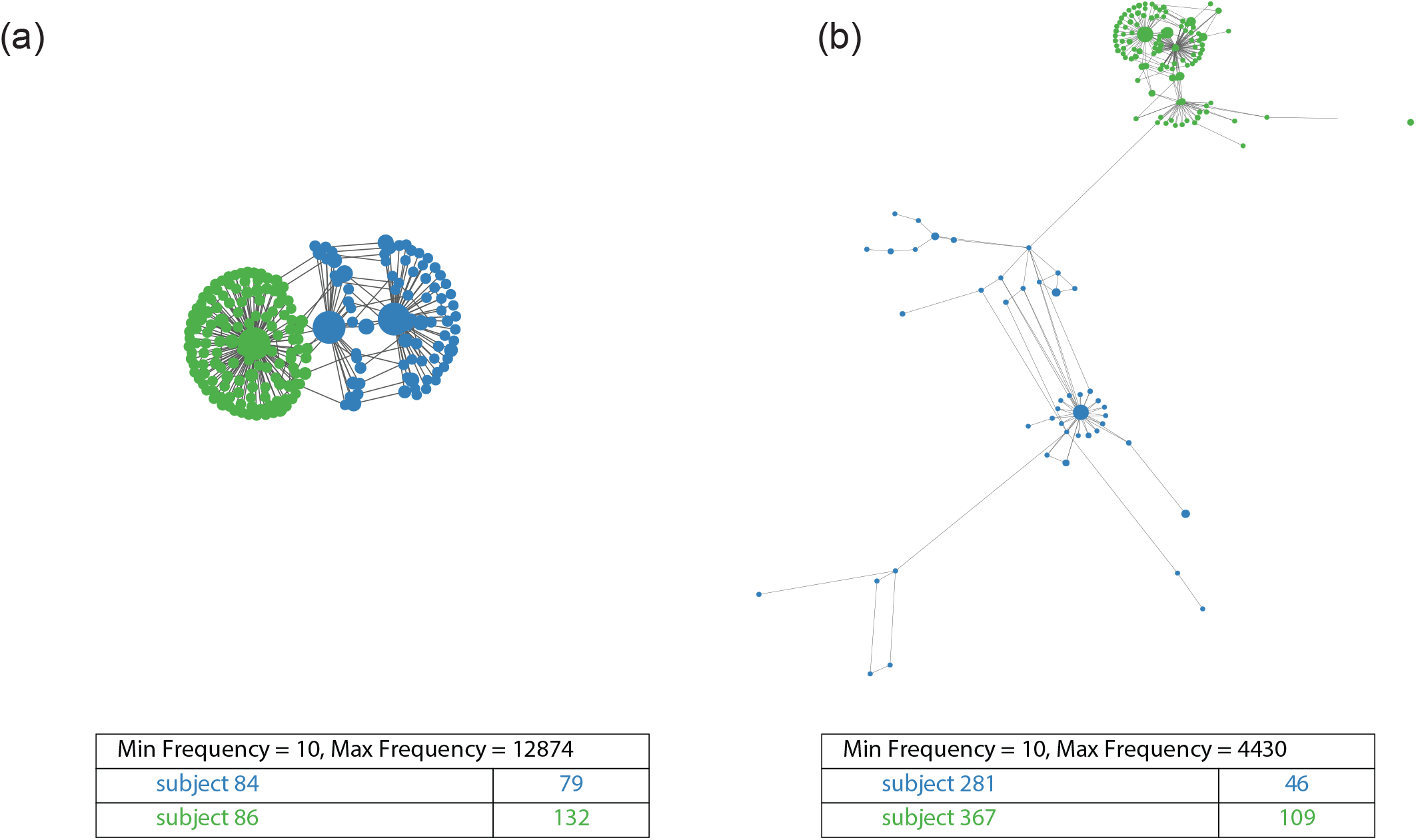
Intra-host genetic variation of representative transmission clusters highlighting the genetic relatedness of distinct variants. K-step network contains all possible minimum spanning trees and allows the efficient visualization of genetic relatedness among all intra-host HVR1 variants for **(a)** Subjects 84 and 86 **(b)** Subjecta 281 and 367. Each node represents an HCV sequence and the color of the node corresponds to the sample of origin: green if it was only found in the first sample and blue if it was only found in the second sample. The node size is based on frequency of the HVR1 variant and edge length is proportional to the modified Hamming distance (does not count positions with insertions or deletions as differences).

## DISCUSSION

This study is the first to link state-wide public health surveillance to HCV transmission clusters identified by GHOST. Among 459 individuals who tested HCV-positive in public health settings in Wisconsin from 2016 to 2017, 424 had sufficient plasma remaining after standard antibody and RNA testing procedures with 379 (89.4%) successfully analyzed by GHOST. Of these, 126 (33%) were detected among 42 HCV transmission clusters. The majority of clusters were formed of dyads, with larger clusters linking multiple sequences less abundant in the populations tested. This rate of clustering is comparable to that found in Vancouver and Baltimore, which found 31% and 46% of PWIDs cluster, respectively *(25,26)*. However, an important distinction between these studies and ours is the geographical region sampled. Whereas these prior studies only included PWIDs from their respective metropolitan areas, this study included both urban and rural populations. We found a higher clustering rate in the rural catchment area, and rural individuals that clustered were of younger age, a finding that aligns with the current literature describing the particular burden of HCV on young persons in rural communities *(27,28)*. Moreover, these data highlight that the increasing rurality of opioid injection and HCV transmission among young PWIDs could be better supported by the expansion of molecular-based surveillance strategies to reduce transmission. The availability of transmission networks would allow for the underlying contact network structure to be targeted such that individuals that have a high centraility within a network have a much greater contribution to infection than peripheral nodes. This type of network-based disruption strategy has been demonstrated to produce the greatest reduction in incidence compared to randomly targeted prevention strategies *(29)*.

The CDC estimates that approximately half of all HCV-infected persons are unaware of their infection status *(30)*. Using molecular epidemiological methods to investigate the transmission of infectious diseases has many advantages over traditional contact tracing methods, in which data collection is often time-intensive and results may be subject to recall and social desirability biases. Contact tracing among people who engage in illegal activity is especially challenging as individuals are often reluctant to disclose injecting behaviors or name injecting partners because of stigma or fear of criminal repercussions *(31)*. GHOST overcomes these limitations by automatically interpreting biological data in a timely manner (within hours). Near real-time monitoring systems already exists for HIV-1 and the application of an automated phylogenetic system has been used to detect and rapidly respond to an outbreak of transmitted drug resistance in Vancouver *(32)*. Thus, a greater investment in viral surveillance would allow for enhanced outbreak control responses and analytical frameworks such as those implemented in GHOST and could be readily applied to assist in understanding HCV transmission dynamics in the U.S. As modeling studies have demonostrated that elimination of HCV can be achieved through scaling up and targeting treatment *(33,34)*, a concept know as ‘treatment-as-prevention’ often used in HIV research *(35,36)*, conducting routine molecular surveillance may also advance HCV prevention efforts by facilitating the ability of health departments to efficiently allocate limited resources to target and treat members in large clusters. Furthermore, it could be used to aid in discriminating recent versus long-established introduction of infections in high-risk communities.

This study has several limitations. First, HCV testing and surveillance challenges make it difficult to identify a complete cohort of HCV-infected PWIDs. Individuals not included in this analysis include those who have never been tested, those tested outside of Wisconsin, and people who were tested in other settings (e.g. primary care) that use commercial or hospital-based laboratories for HCV confirmatory testing. Accordingly, the population studied is not fully representative of the Wisconsin general population. However, our results do provide a credible picture of the HCV epidemic across public health settings throughout rural and urban Wisconsin. Second, the association we found between clustering and younger age among rural residents could be due to sampling a larger number of younger persons. Further research is needed examining the role that older HCV-infected individuals play in transmission networks. Third, risk information data are missing for a large proportion of the sample because few agencies routinely collect and report it. Although this limited the types of analyses we could conduct, it also demonstrates where data collection from different types of public health agencies can be improved to better understand the characteristics of HCV transmission clusters on a state level. Lastly, phylogenetic clustering alone cannot directly assert whether transmission occurred directly or indirectly via unsampled individuals and can be subjected to biases *(37)*.

In conclusion, we provided a snapshot of the HCV epidemic in rural and urban Wisconsin between 2016-2017. Additional research to understand the dynamics of HCV transmission is urgently needed particularly in rural communities affected by the opioid crisis. Incorporating GHOST into routine laboratory surveillance practices may enhance the ability of public health departments to detect individuals contributing most to HCV transmission in their communities and prompt more efficient interventions.

## Data Availability

The data that support these findings will be available upon request.

## Prior Presentations

The methods described in this paper, along with preliminary results from the first 231 specimens analyzed, were presented in poster format at the 8^th^ International Conference on Hepatitis Care in Substance Users in Montreal, Canada on September 11-13, 2019.

## Biographical Sketch

Karli R Hochstatter, PhD, MPH is a Post-Doctoral Research Fellow at the Columbia University School of Social Work in New York and an Honorary Fellow at the University of Wisconsin School of Medicine and Public Health in Madison. Her research focuses on preventing HIV and HCV transmission and improving screening and treatment uptake among populations disproportionately burdened by substance use disorders, with a special focus on people who inject drugs and criminal justice-involved adults.

Damien C. Tully PhD is a Assistant Professor in Epidemiology, Biostatistics and Bioinformatics at the London School of Hygiene and Tropical Medicine. His primary research interests include the evolution and molecular epidemiology of RNA viruses.

## REFERENCES

1. Hofmeister MG, Rosenthal EM, Barker LK, Rosenberg ES, Barranco MA, Hall EW, et al. Estimating Prevalence of Hepatitis C Virus Infection in the United States, 2013-2016. Hepatology. 2019 Mar;69(3):1020–31.

2. Ly KN, Hughes EM, Jiles RB, Holmberg SD. Rising Mortality Associated With Hepatitis C Virus in the United States, 2003–2013. Clinical Infectious Diseases. 2016 May 15;62(10):1287–8.

3. Zibbell JE, Asher AK, Patel RC, Kupronis B, Iqbal K, Ward JW, et al. Increases in Acute Hepatitis C Virus Infection Related to a Growing Opioid Epidemic and Associated Injection Drug Use, United States, 2004 to 2014. American Journal of Public Health. 2018 Feb;108(2):175–81.

4. Gardy JL, Johnston JC, Sui SJH, Cook VJ, Shah L, Brodkin E, et al. Whole-Genome Sequencing and Social-Network Analysis of a Tuberculosis Outbreak. New England Journal of Medicine. 2011 Feb 24;364(8):730–9.

5. Little SJ, Kosakovsky Pond SL, Anderson CM, Young JA, Wertheim JO, Mehta SR, et al. Using HIV Networks to Inform Real Time Prevention Interventions. Harrigan PR, editor. PLoS ONE. 2014 Jun 5;9(6):e98443.

6. Poon AFY, Gustafson R, Daly P, Zerr L, Demlow SE, Wong J, et al. Near real-time monitoring of HIV transmission hotspots from routine HIV genotyping: an implementation case study. The Lancet HIV. 2016 May;3(5):e231–8.

7. Gardy JL, Loman NJ. Towards a genomics-informed, real-time, global pathogen surveillance system. Nature Reviews Genetics. 2017 Nov 13;19(1):9–20.

8. Fauci AS, Redfield RR, Sigounas G, Weahkee MD, Giroir BP. Ending the HIV Epidemic. JAMA. 2019 Mar 5;321(9):844.

9. Longmire AG, Sims S, Rytsareva I, Campo DS, Skums P, Dimitrova Z, et al. GHOST: global hepatitis outbreak and surveillance technology. BMC Genomics. 2017 Dec 6;18(S10):916.

10. Vivolo-Kantor AM, Seth P, Gladden RM, Mattson CL, Baldwin GT, Kite-Powell A, et al. Vital Signs: Trends in Emergency Department Visits for Suspected Opioid Overdoses - United States, July 2016-September 2017. MMWR Morbidity and mortality weekly report. 2018 Mar 9;67(9):279–85.

11. Wisconsin Department of Health Service, Wisconsin Epidemiological Profile on Alcohol and Other Drugs, 2016 (P-45718-16).

12. Centers for Disease Control and Prevention (CDC). Surveillance for Viral Hepatitis – United States, 2016. 2018.

13. Tully DC, Hjerrild S, Leutscher PD, Renvillard SG, Ogilvie CB, Bean DJ, et al. Deep sequencing of hepatitis C virus reveals genetic compartmentalization in cerebrospinal fluid from cognitively impaired patients. Liver International. 2016;36(10).

14. Campo DS, Xia G-L, Dimitrova Z, Lin Y, Forbi JC, Ganova-Raeva L, et al. Accurate Genetic Detection of Hepatitis C Virus Transmissions in Outbreak Settings. Journal of Infectious Diseases. 2016 Mar 15;213(6):957–65.

15. Tully DC, Ogilvie CB, Batorsky RE, Bean DJ, Power KA, Ghebremichael M, et al. Differences in the Selection Bottleneck between Modes of Sexual Transmission Influence the Genetic Composition of the HIV-1 Founder Virus. PLoS Pathogens. 2016;12(5).

16. Henn MR, Boutwell CL, Charlebois P, Lennon NJ, Power KA, Macalalad AR, et al. Whole genome deep sequencing of HIV-1 reveals the impact of early minor variants upon immune recognition during acute infection. PLoS Pathogens. 2012;8(3).

17. Hedegaard DL, Tully DC, Rowe IA, Reynolds GM, Bean DJ, Hu K, et al. High resolution sequencing of hepatitis C virus reveals limited intra-hepatic compartmentalization in end-stage liver disease. Journal of Hepatology. 2017;66(1).

18. Xu H, Luo X, Qian J, Pang X, Song J, Qian G, et al. FastUniq: a fast de novo duplicates removal tool for paired short reads. Doucet D, editor. PloS one. 2012 Dec 20;7(12):e52249.

19. Bolger AM, Lohse M, Usadel B. Trimmomatic: a flexible trimmer for Illumina sequence data. Bioinformatics. 2014 Aug 1;30(15):2114–20.

20. Yang X, Charlebois P, Gnerre S, Coole MG, Lennon NJ, Levin JZ, et al. De novo assembly of highly diverse viral populations. BMC Genomics. 2012 Sep 13;13(1):475.

21. Tamura K, Stecher G, Peterson D, Filipski A, Kumar S. MEGA6: Molecular Evolutionary Genetics Analysis Version 6.0. Molecular Biology and Evolution. 2013 Dec;30(12):2725–9.

22. Nguyen L-T, Schmidt HA, von Haeseler A, Minh BQ. IQ-TREE: A Fast and Effective Stochastic Algorithm for Estimating Maximum-Likelihood Phylogenies. Molecular Biology and Evolution. 2015 Jan;32(1):268–74.

23. Hoang DT, Chernomor O, von Haeseler A, Minh BQ, Vinh LS. UFBoot2: Improving the Ultrafast Bootstrap Approximation. Molecular Biology and Evolution. 2018 Feb 1;35(2):518–22.

24. Sims S, Longmire AG, Campo DS, Ramachandran S, Medrzycki M, Ganova-Raeva L, et al. Automated quality control for a molecular surveillance system. BMC Bioinformatics. 2018 Oct 22;19(S11):358.

25. Jacka, Applegate T, Krajden M, Olmstead A, Harrigan PR, Marshall BDL, et al. Phylogenetic clustering of hepatitis C virus among people who inject drugs in Vancouver, Canada. Hepatology. 2014 Nov;60(5):1571–80.

26. Hackman J, Falade-Nwulia O, Patel EU, Mehta SH, Kirk GD, Astemborski J, et al. Correlates of hepatitis C viral clustering among people who inject drugs in Baltimore. Infection, Genetics and Evolution. 2020 May 16;77:104078.

27. Suryaprasad AG, White JZ, Xu F, Eichler B-A, Hamilton J, Patel A, et al. Emerging Epidemic of Hepatitis C Virus Infections Among Young Nonurban Persons Who Inject Drugs in the United States, 2006–2012. Clinical Infectious Diseases. 2014 Nov 15;59(10):1411–9.

28. Van Handel MM, Rose CE, Hallisey EJ, Kolling JL, Zibbell JE, Lewis B, et al. County-Level Vulnerability Assessment for Rapid Dissemination of HIV or HCV Infections among Persons Who Inject Drugs, United States. In: Journal of Acquired Immune Deficiency Syndromes. Lippincott Williams and Wilkins; 2016. p. 323–31.

29. Campo DS, Khudyakov Y. Intelligent Network DisRuption Analysis (INDRA): A targeted strategy for efficient interruption of hepatitis C transmissions. Infection, Genetics and Evolution. 2018 Sep;63:204–15.

30. Centers for Disease Control and Prevention. Hepatitis C Kills More Americans than Any Other Infectious Disease | CDC Online Newsroom | CDC [Internet]. 2016 [cited 2019 Sep 24]. Available from: https://www.cdc.gov/media/releases/2016/p0504-hepc-mortality.html

31. Katzman C, Mateu-Gelabert P, Kapadia SN, Eckhardt BJ. Contact tracing for hepatitis C: The case for novel screening strategies as we strive for viral elimination. The International journal on drug policy. 2019 Oct;72:33–9.

32. Poon AFY, Gustafson R, Daly P, Zerr L, Demlow SE, Wong J, et al. Near real-time monitoring of HIV transmission hotspots from routine HIV genotyping: An implementation case study. The Lancet HIV. 2016 May 1;3(5):e231–8.

33. Mathematical modeling of hepatitis c virus (HCV) prevention among people who inject drugs: A review of the literature and insights for elimination strategies. - Abstract – Europe PMC [Internet]. [cited 2020 Jun 13]. Available from: https://europepmc.org/article/pmc/pmc6522340

34. Heffernan A, Cooke GS, Nayagam S, Thursz M, Hallett TB. Scaling up prevention and treatment towards the elimination of hepatitis C: a global mathematical model. The Lancet. 2019 Mar 30;393(10178):1319–29.

35. Cohen MS, McCauley M, Gamble TR. HIV treatment as prevention and HPTN 052. Curr Opin HIV AIDS. 2012 Mar;7(2):99–105.

36. Cohen J. HIV Treatment as Prevention. Science. 2011 Dec 23;334(6063):1628–1628.

37. Villabona-Arenas ChJ, Hanage WP, Tully DC. Phylogenetic interpretation during outbreaks requires caution. Nature Microbiology. 2020 May 19;1–2.

